# Trends in severe acute malnutrition admissions, characteristics, and treatment outcomes in Malawi from 2011 through 2019

**DOI:** 10.1101/2022.06.17.22276562

**Authors:** Allison I. Daniel, Sylvester Kathumba, Collins Mitambo, Dennis Chasweka, Wieger Voskuijl, Esther Kamanga, Emmie Mbale, Robert H.J. Bandsma, Isabel Potani

**Author notes:** **Corresponding author:** Dr. Allison Daniel, Peter Gilgan Centre for Research and Learning, 686 Bay Street, Toronto, ON M5G 0A4, or 0991514203. RHJB and IP are co-senior authors.

## Abstract

**Background:** Community-based Management of Acute Malnutrition (CMAM) has been successfully implemented across Malawi, yet trends in admissions, characteristics, and treatment outcomes in children with severe acute malnutrition (SAM) have not been examined. The objective was therefore to investigate trends in admissions, characteristics including percentage of children with SAM with HIV and oedema, and treatment outcomes across the decade following implementation of CMAM.

**Methods:** This research involved a retrospective analysis of existing data routinely collected across Malawi by the Ministry of Health between 2011 and 2019.

**Results:** These data showed an increase in outpatient therapeutic feeding (OTP) admissions from 30323 children in 2011 to 37655 in 2019 (p=0.045). However, a significant decrease in nutritional rehabilitation unit (NRU) admissions was observed, from 11389 annual admissions in 2011 to 6271 in 2019 (p=0.006). In children identified with SAM, the percentage with oedema decreased in OTPs with an average annual rate of reduction (AARR) of 5.6% (p=0.001) and by 26.2% in NRUs in this timeframe with an AARR of 8.5% (p<0.001). There was a decrease in the percentage of children with SAM who had HIV over time in OTPs with an AARR of 16.1% (p=0.001). HIV rates also decreased in NRUs with an AARR of 7.2% (p=0.4), but this difference was not significant. Death rates decreased in OTPs with an AARR of 6.0% (p=0.01). Mortality rates did not change in NRUs over time with an AARR of 0.9% (p=0.5) with the NRU mortality rate in 2019 being 11.0%.

**Conclusions:** These trends indicate that there has been an increase in OTP admissions and a corresponding decrease in NRU admissions. There have been decreases in the percentage of children with oedematous SAM in OTPs and in NRUs and with HIV in OTPs. Children remain at high risk of mortality in NRUs.

**Competing interests:** none to declare

## Introduction

The Community-based Management of Acute Malnutrition (CMAM) approach, which was endorsed by the World Health Organization, UNICEF, and World Food Programme in 2007, drastically changed the way that acute malnutrition is managed (World Health Organization, 2007). Severe acute malnutrition (SAM) is defined as severe wasting (weight-for-height z-scores (WHZ) below -3 SD or mid-upper arm circumference (MUAC) below 115 mm) and/or oedematous malnutrition (bilateral pitting oedema). Children with SAM and acute illnesses, loss of appetite, or other medical complications require admission to nutritional rehabilitation units (NRUs) for clinical care and nutritional support. Children with SAM in the absence of complications are treated in outpatient therapeutic feeding programs (OTP), including those discharged from NRUs. Children with moderate wasting, otherwise known as moderate acute malnutrition (MAM) (WHZ between -3 SD and -2 SD or MUAC between 115mm and 125mm), are commonly managed within supplementary feeding programs (SFP) (World Health Organization, 2009).

Malawi has seen a downward trend in the prevalence of wasting from 7% in 2000 to 3% in 2015 (National Statistical Office/Malawi ICF, 2017). CMAM was first established in 2002 in Malawi as a pilot program which was then implemented nationally in 2006 to manage children with acute malnutrition as part of the national nutrition policy and strategy in Malawi (Concern Worldwide, 2013; Kathumba, 2012; Malawi Ministry of Health, 2016). Scale-up continued until all 28 districts in the country implemented CMAM programs by 2009 (Kathumba, 2012). There are 104 operational NRUs in Malawi for inpatient treatment of SAM and over 620 OTP centers (Kouam, 2016). In 2019, coverage for SAM treatment was 67.3% (38610 children reached out of a target 75% of 76509 caseloads).

While CMAM has been successfully implemented in Malawi, there has been little examination of trends in admissions, characteristics, and treatment outcomes since its inception in the country which would aid in resource-allocation within the CMAM approach and to characterize the population of children with SAM in different treatment settings. The main objective of this analysis was therefore to examine trends in SAM admissions within the last decade prior to the COVID-19 pandemic, from 2011 to 2019, at CMAM sites across the country based on pre-existing data collected by the Malawi Ministry of Health. Another important aim was to understand trends in characteristics of children with SAM and trends in treatment outcomes including mortality in children with SAM.

## Methods

This research involved a retrospective analysis of existing CMAM data that were routinely collected across Malawi by the Ministry of Health between 2011 and 2019 at up to 104 NRUs and 623 OTPs. Within each site, health workers or other personnel record the number of children admitted and discharged and a supervisor compiles a report at the end of each month which is sent to the District Health Office. The mean and median number of NRUs and OTPs reporting each month across the different years is summarized in Table 1. The number of units reporting was not documented prior to 2016, apart from 2011 in which the mean number of NRUs was 98.7 and median was 100. Note that data during the COVID-19 pandemic were not included in this analysis because they were not collected consistently throughout this time and there were disruptions in accessing routine health and nutrition services.

**Table 1.** Outpatient therapeutic feeding programs and nutritional rehabilitation units reporting each month between 2016 and 2019.

| **Year** | **OTP** |  | **NRU** |  |
| --- | --- | --- | --- | --- |
|  | **Mean** | **Median** | **Mean** | **Median** |
| 2016<sup>1</sup> | 595.3<sup>a</sup> | 600<sup>1</sup> | 103 | 103 |
| 2017 | 613.4 | 613 | 103.9 | 104 |
| 2018 | 619.4 | 619 | 103.9 | 104 |
| 2019 | 623 | 623 | 104 | 104 |

NRU: nutritional rehabilitation unit; OTP: outpatient therapeutic feeding program.
<sup>a</sup>Based on the number of centres reported for nine of 12 months in 2016.

Aggregate data for each district were obtained from the Malawi Ministry of Health. All variables with sufficient data across multiple years were considered for the purpose of this analysis. Data extracted included the number of SFP, NRU, and OTP admissions, characteristics, and treatment outcomes including mortality. The characteristics included the percentage of children identified with SAM who had oedema out of total admissions (data available from 2011 to 2018), percentage of children with SAM with HIV versus children with SAM without HIV (data available from 2012 to 2017), and gender (data available to 2011 and 2018). In 2011, severe wasting was identified by weight-for-height below 70% and/or MUAC below 110mm; from 2012 onwards, WHZ <-3 SD and/or MUAC below 115mm were used. The NRU and OTP data up to 2018 include children up to five years of age, while children above five years are also included in the 2019 data. The 2019 data also only capture new admissions, while data from all other years reflect total admissions. Monthly admissions between the different years were examined based on data that were available from 2011 to 2018.

All data were analyzed using statistical software Stata 16 (StataCorp LP, College Station, Texas, USA) (StataCorp, 2019). Annual trends in admissions and sources of admissions to NRUs and OTP, the percentage of children with SAM who had oedema out of total admissions, the percentage of children with HIV versus children without HIV disaggregated by CMAM setting, the percentage of children with SAM who were female versus male disaggregated by CMAM setting, and treatment outcomes disaggregated by CMAM setting were evaluated using a linear-by-linear trend test with percentage point changes presented. These annual trends, as well as SAM admissions by month, are also shown visually. The average annual rate of reduction (AARR) was also calculated for the above trends in percentages where applicable.

### Ethics Statement

Ethical approval for this analysis was obtained from the Malawi National Health Sciences Research Committee (Protocol #20/01/2459) in Lilongwe, Malawi. The information used in the analysis was routinely collected programmatic data; no individual patient information nor identifying information were collected and therefore informed consent was not required.

## Results

### Admissions for acute malnutrition

SFP admissions for management of moderate wasting rose from 53446 to 147696 children representing a percentage point increase of 176.3% between 2011 and 2019 (p=0.01). Total SAM admissions to both OTPs and NRUs, which include new admissions and readmissions, went from 41712 to 49167, which was a 17.9% percentage point increase, but this trend was not significant (p=0.06) (Figure 1). New SAM admissions to OTPs and NRUs specifically increased by 24.0% percentage points, from 35416 to 43926 (p=0.04).

**Figure 1.**
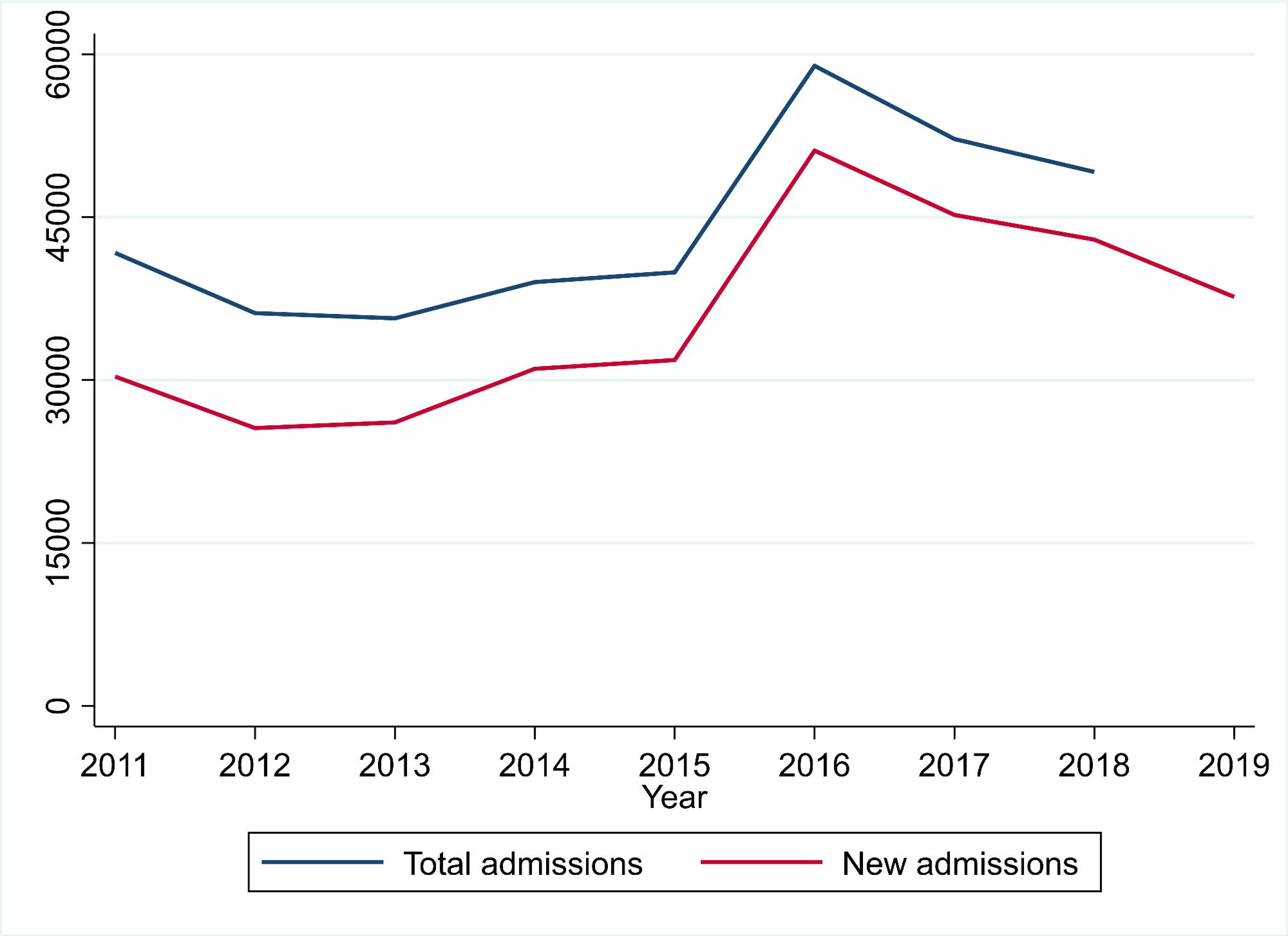
Severe acute malnutrition admissions between 2011 and 2019. Total and new severe acute malnutrition admissions by year at all nutritional rehabilitation units and outpatient therapeutic feeding programs across Malawi.

There was a significant positive trend in OTP admissions specifically (p=0.045) from 30323 to 37655 children with SAM which was a 24.2% percentage point increase (Figure 2). NRU admissions decreased significantly (p=0.006) from 11389 to 6271 between 2011 and 2019, by 44.9% percentage points (Figure 3). With regards to monthly SAM admissions between 2011 and 2018 at NRUs and OTP, the highest numbers were generally seen in the first three months of the year (Figure 4).

**Figure 2.**
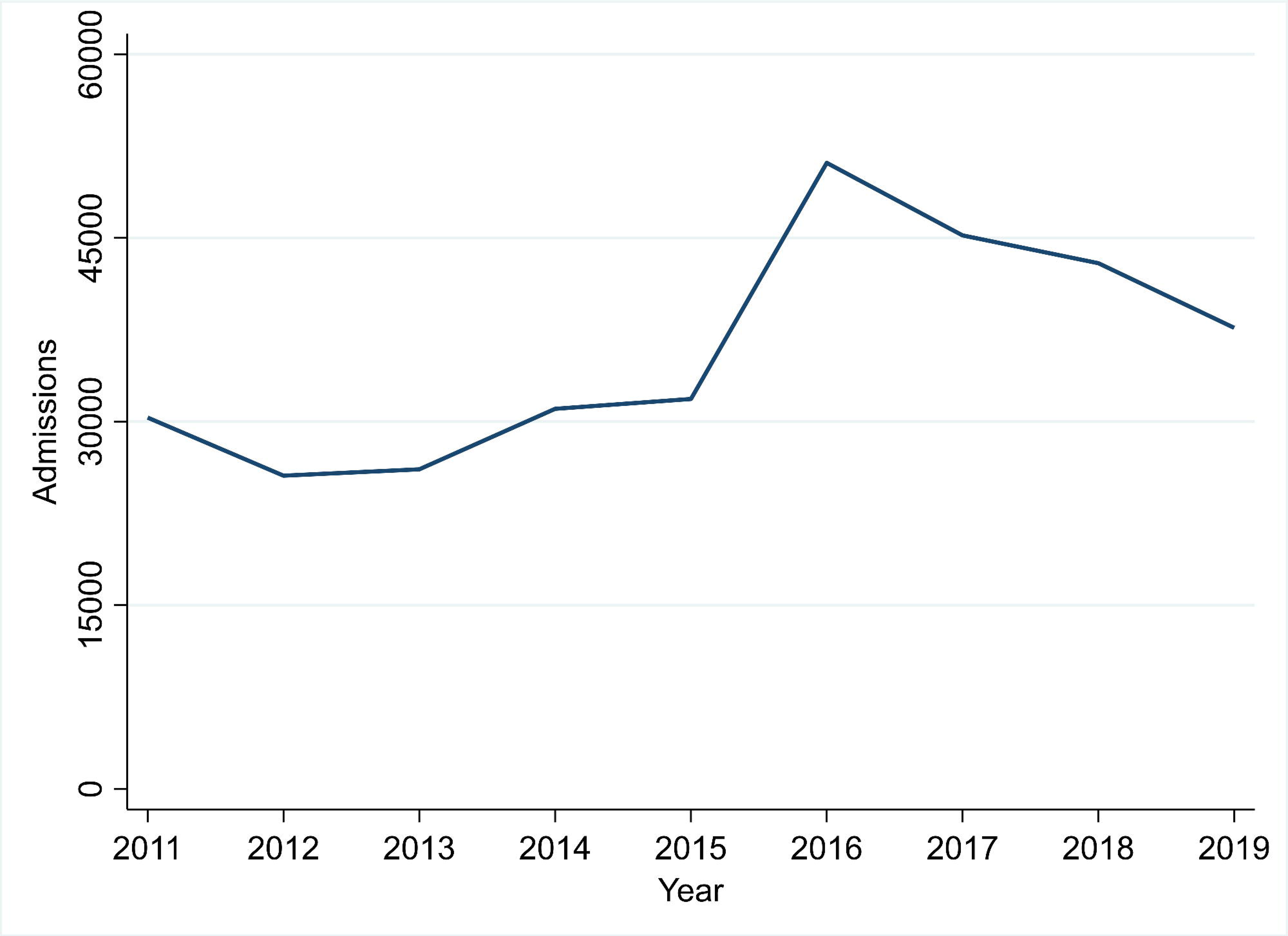
Outpatient therapeutic feeding program admissions between 2011 and 2019. Severe acute malnutrition admissions by year at all outpatient therapeutic feeding programs across Malawi. The 2019 data only include new admissions while data from all other years reflect total admissions.

**Figure 3.**
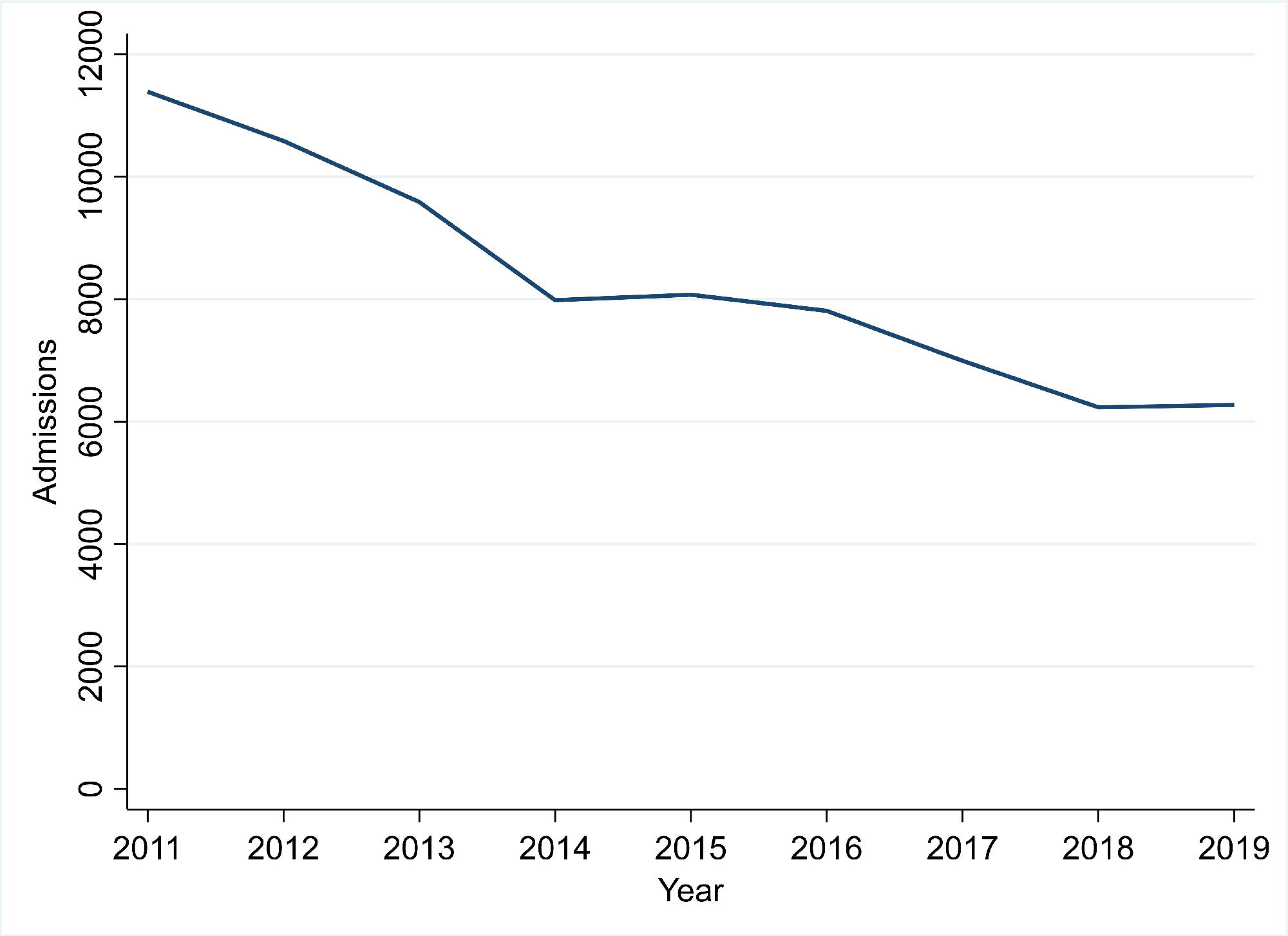
Nutritional rehabilitation unit admissions between 2011 and 2019. Severe acute malnutrition admissions by year at all nutritional rehabilitation units across Malawi. The 2019 data only include new admissions while data from all other years reflect total admissions.

**Figure 4.**
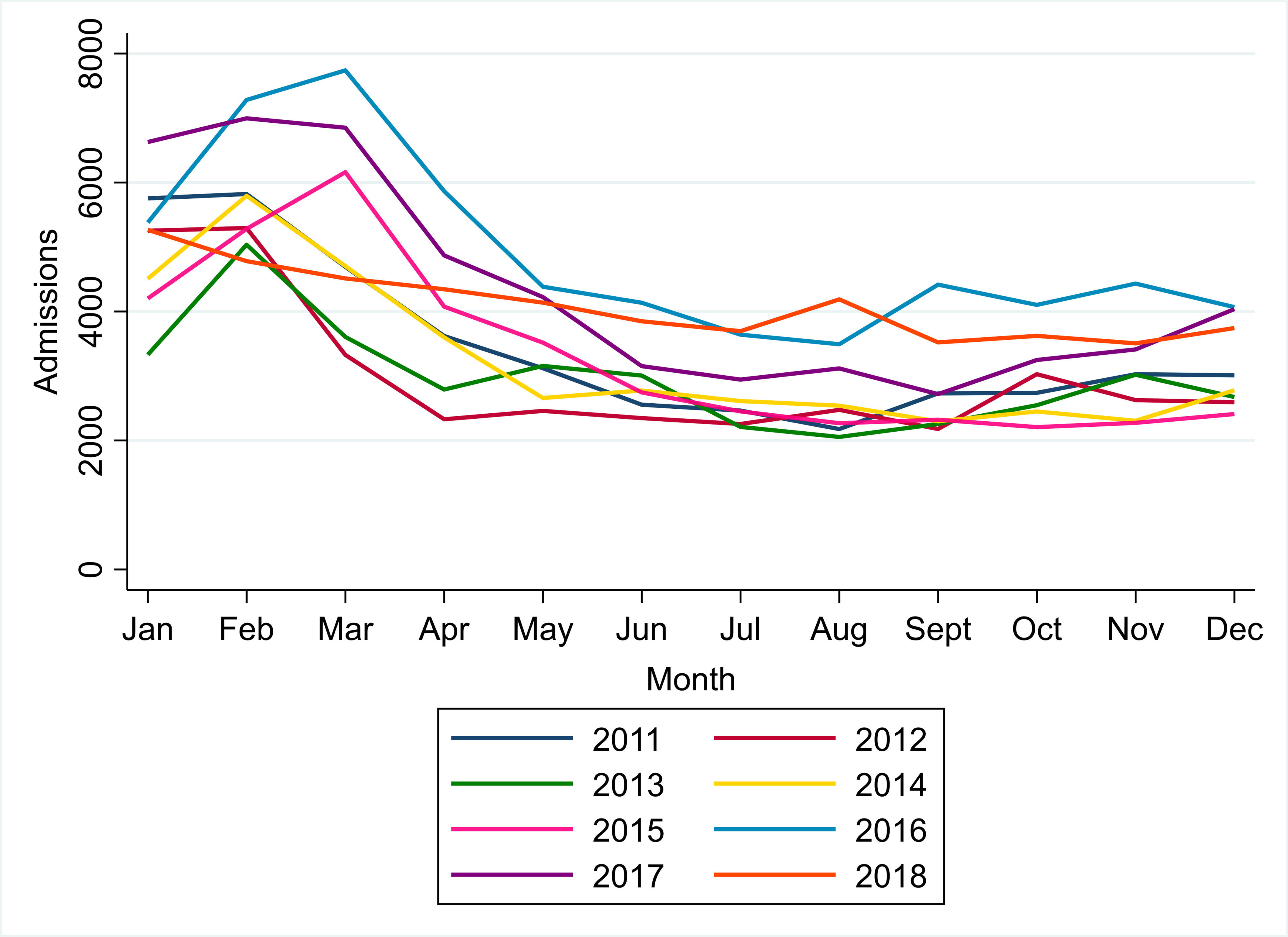
Severe acute malnutrition admissions by month between 2011 and 2018. Total severe acute malnutrition admissions by month at all outpatient therapeutic feeding programs and nutritional rehabilitation units across Malawi.

There was no significant decrease in the percentage of children with SAM admitted to NRUs who were transferred from hospital (p=0.4) and there was an AARR of 7.6% (95% CI: - 12.6%, 24.2%) (Table 2). There was no significant decrease in the percentage of children with SAM transferred from other OTPs to NRUs (p=0.3) with an AARR of 2.0% (95% CI: -3.4%, 7.2%). There was a reduction in the rates of returned defaulters in OTP (p=0.03) with an AARR of 11.4% (95% CI: 2.9%, 19.2%). There was also a significant decline in transfer from NRUs to OTP (p=0.01) with an AARR of 18.1% (95% CI: 13.6%, 22.3%) or from other OTPs (p=0.01) with an AARR of 11.5% (95% CI: 9.2%, 13.7%). There was an AARR of -20.5% (95% CI: - 46.6%, 0.9%) in transfers from SFP to OTP.

**Table 2.** Sources of admissions to outpatient therapeutic feeding programs and nutritional rehabilitation units between 2011 and 2018.

| Year | NRU |  | OTP |  |  |  |
| --- | --- | --- | --- | --- | --- | --- |
|  | Transfer from hospital | Transfer from other OTP | Returned defaulter | Transfer from NRU | Transfer from other OTP | Transfer from SFP |
| 2011 | 0.8% | 3.8% | 1.8% | 17.4% | 2.9% | 0.4% |
| 2012 | 0.7% | 2.5% | 1.5% | 14.4% | 2.4% | 1.8% |
| 2013 | 2.6% | 3.6% | 1.3% | 13.0% | 2.3% | 1.9% |
| 2014 | 1.3% | 3.1% | 1.6% | 9.0% | 1.7% | 2.6% |
| 2015 | 1.2% | 3.4% | 1.6% | 9.6% | 1.6% | 3.0% |
| 2016 | 1.0% | 2.8% | 1.4% | 5.1% | 1.5% | 3.0% |
| 2017 | 0.5% | 3.1% | 0.7% | 5.3% | 1.4% | 2.5% |
| 2018 | 0.6% | 2.8% | 0.7% | 4.8% | 1.2% | 2.4% |
| **Linear-by-linear test** | p=0.4 | p=0.3 | p=0.03 | p=0.01 | p=0.01 | p=0.054 |
| **AARR** | 7.6% (95% CI: -12.6%, 24.2%; p=0.04) | 2.0% (95% CI: -3.4%, 7.2%; p=0.4) | 11.3% (95% CI: 2.9%, 19.2%; p=0.02) | 18.1% (95% CI: 13.6%, 22.3%; p<0.001) | 11.5% (95% CI: 9.2%, 13.7%; p<0.001) | -20.5% (95% CI: -46.6%, 0.9%; p=0.06) |

AARR: average annual rate of reduction; NRU: nutritional rehabilitation unit; OTP: outpatient therapeutic feeding program; SFP: supplementary feeding program.

### Characteristics of children with severe acute malnutrition

In children identified with SAM, the percentage of those with oedema went down over time, from 46.3% to 33.7% (p=0.02) in OTP with an AARR of 5.6% (95% CI: 3.3%, 7.9%) between 2011 and 2018 (Figure 5, Figure 6A) and from 62.8% to 36.6% (p=0.01) in NRUs with an AARR of 8.5% (95% CI: 6.3%, 10.5%) between 2011 and 2018 (Figure 5, Figure 6B).

**Figure 5.**
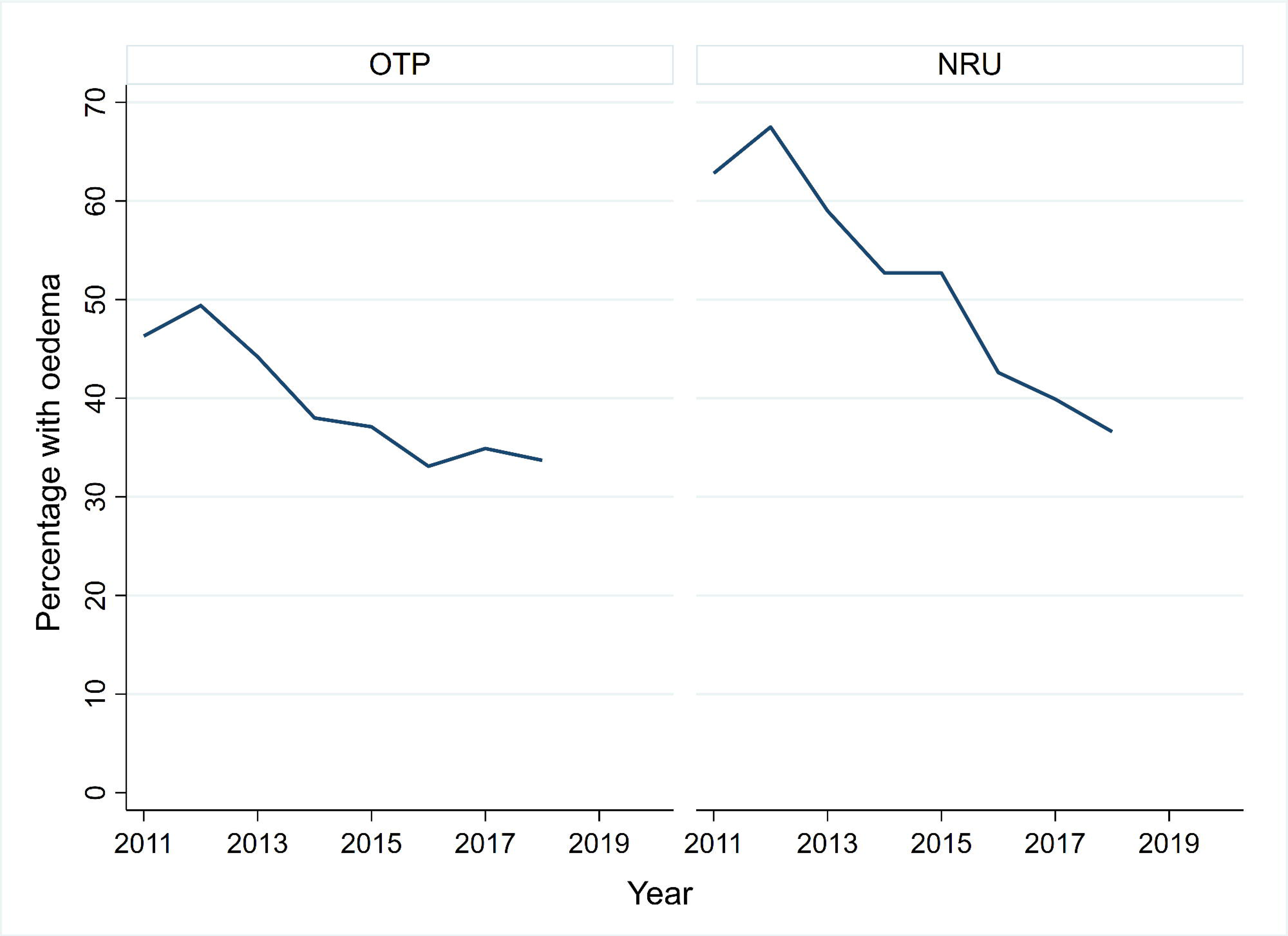
**Trends in the percentage of children with severe acute malnutrition who had oedema in outpatient therapeutic feeding programs (A) and nutritional rehabilitation units (B) between 2011 and 2018.**

**Figure 6A.**
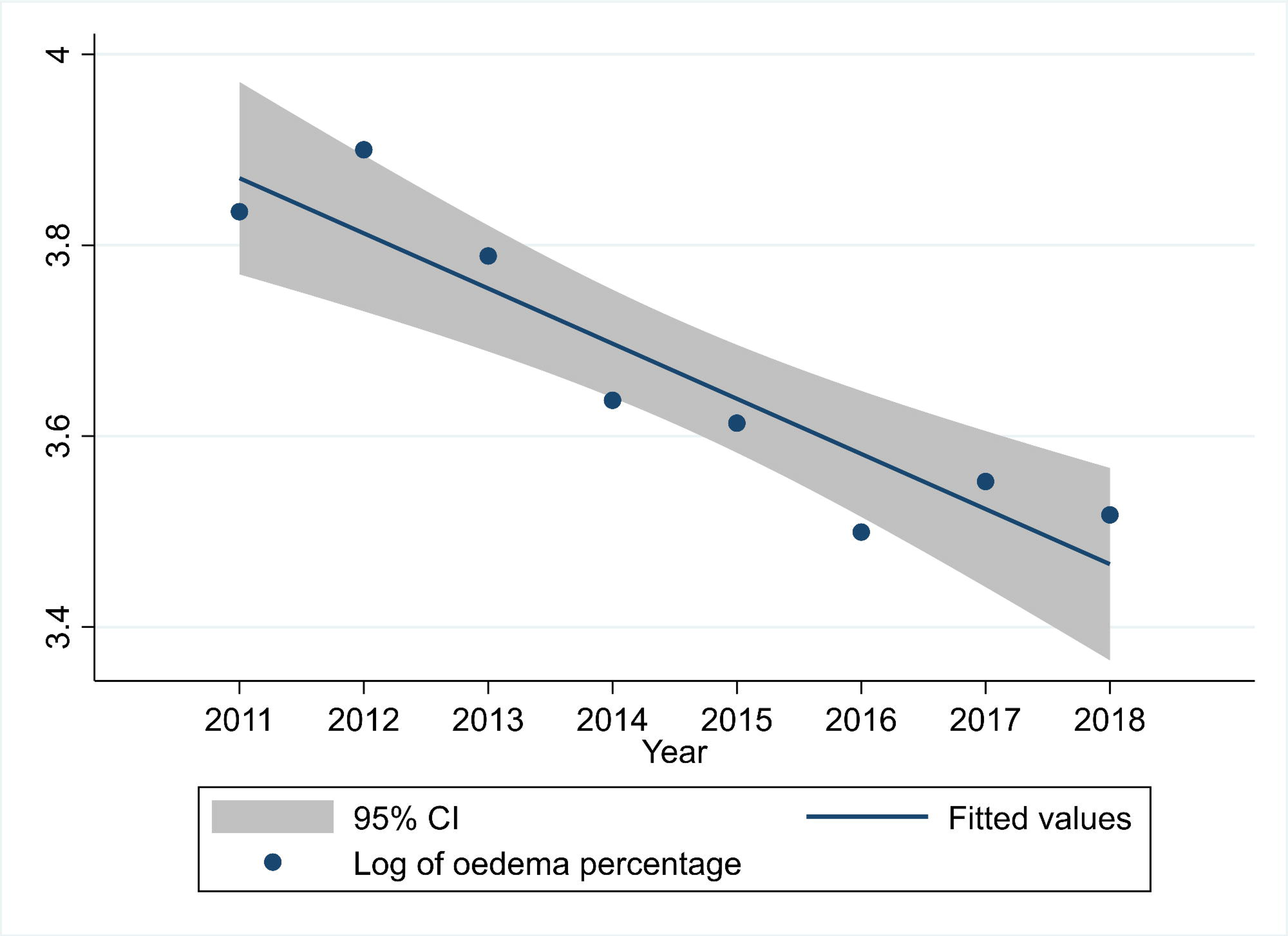
**Average annual rate of reduction in the percentage of children with severe acute malnutrition who had oedema in outpatient therapeutic feeding programs between 2011 and 2018.**

**Figure 6B.**
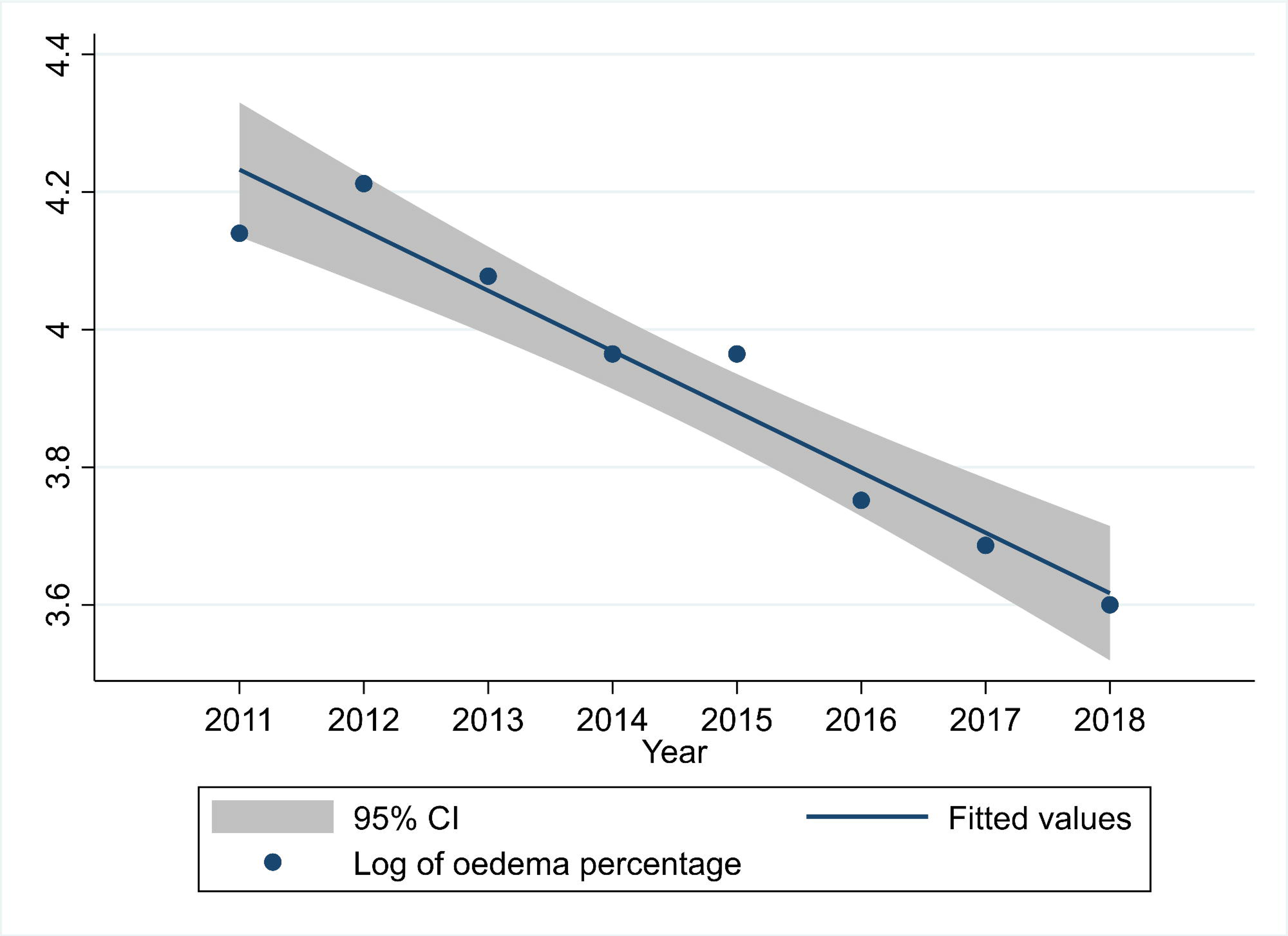
**Average annual rate of reduction in the percentage of children with severe acute malnutrition who had oedema in nutritional rehabilitation units between 2011 and 2018.**

The percentage of children with SAM with HIV in OTP also declined between 2012 and 2017, from 16.5% to 7.1% (p=0.03) with an AARR of 16.1% (95% CI: 11.7%, 20.2%) (Figure 7, Figure 8A). There was no significant trend in HIV percentage in children with SAM admitted to NRUs (p=0.06), although there was a significant AARR of 7.2% (95% CI: 1.4%, 12.7%) (Figure 7, Figure 8B).

**Figure 7.**
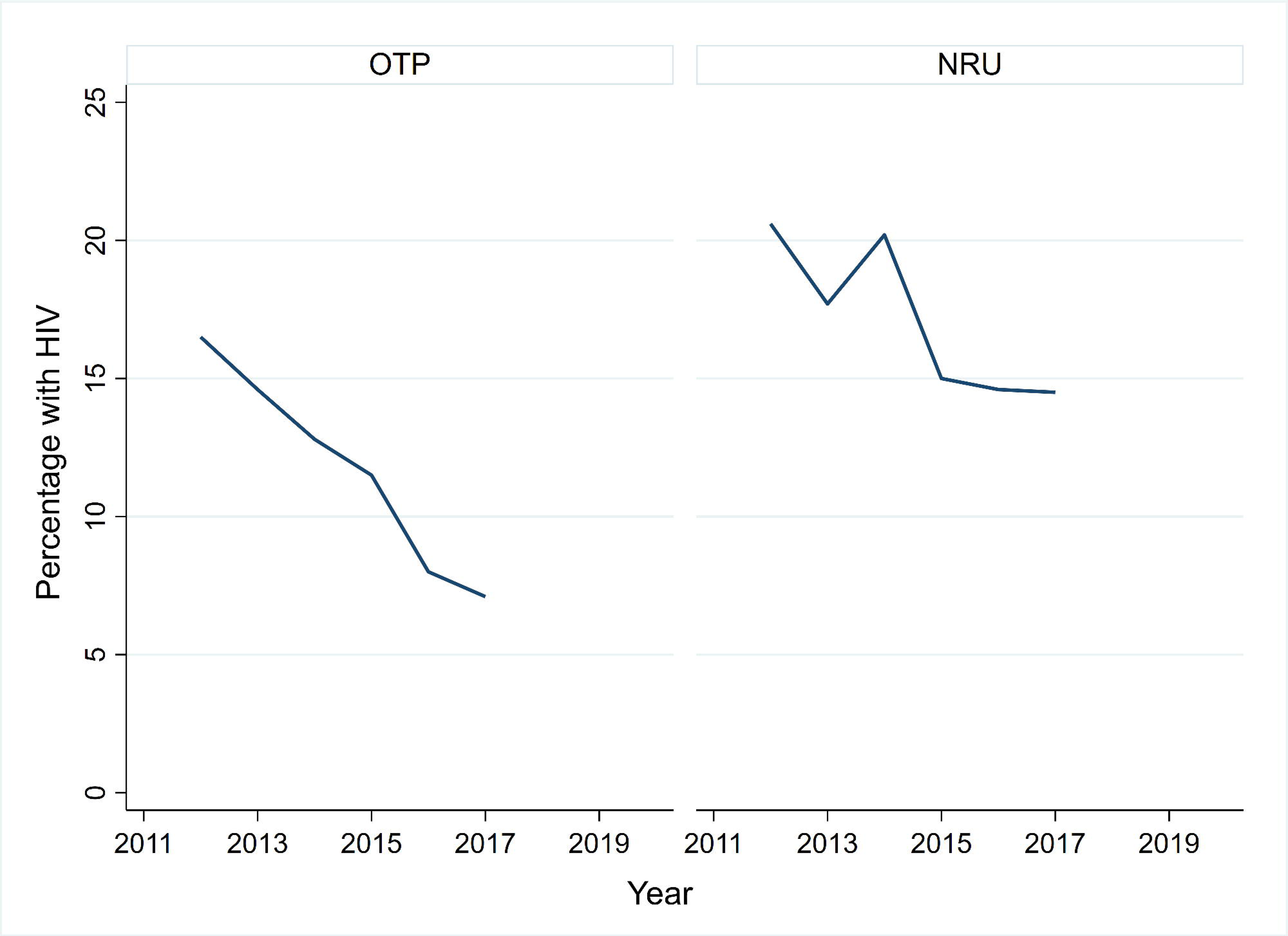
Trends in the percentage of children with severe acute malnutrition who were HIV reactive in outpatient therapeutic feeding programs (A) and nutritional rehabilitation units (B) between 2012 and 2017. HIV: human immunodeficiency virus

**Figure 8A.**
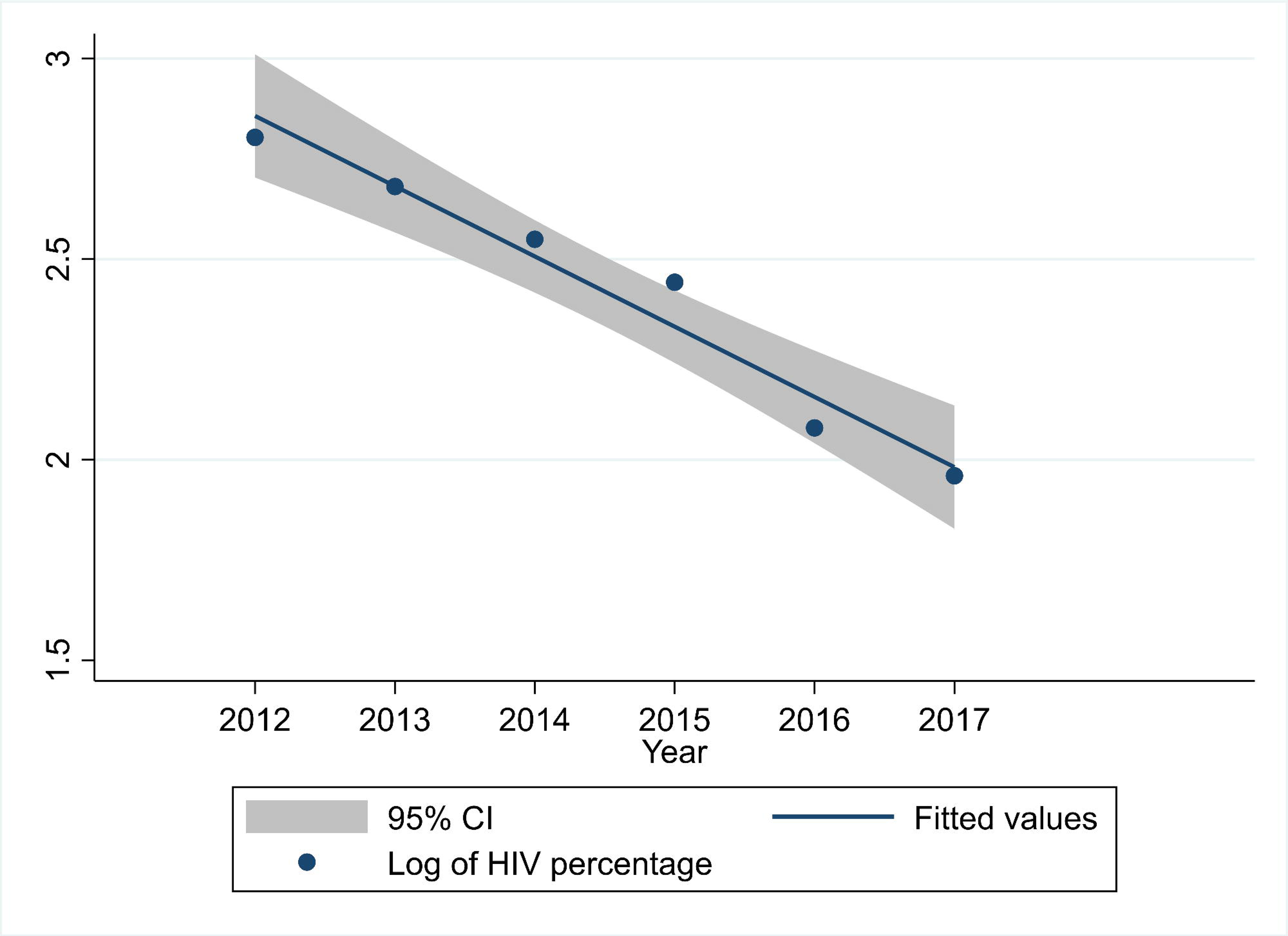
Average annual rate of reduction in the percentage of children with severe acute malnutrition who had HIV in outpatient therapeutic feeding programs between 2012 and 2017. HIV: human immunodeficiency virus

**Figure 8B.**
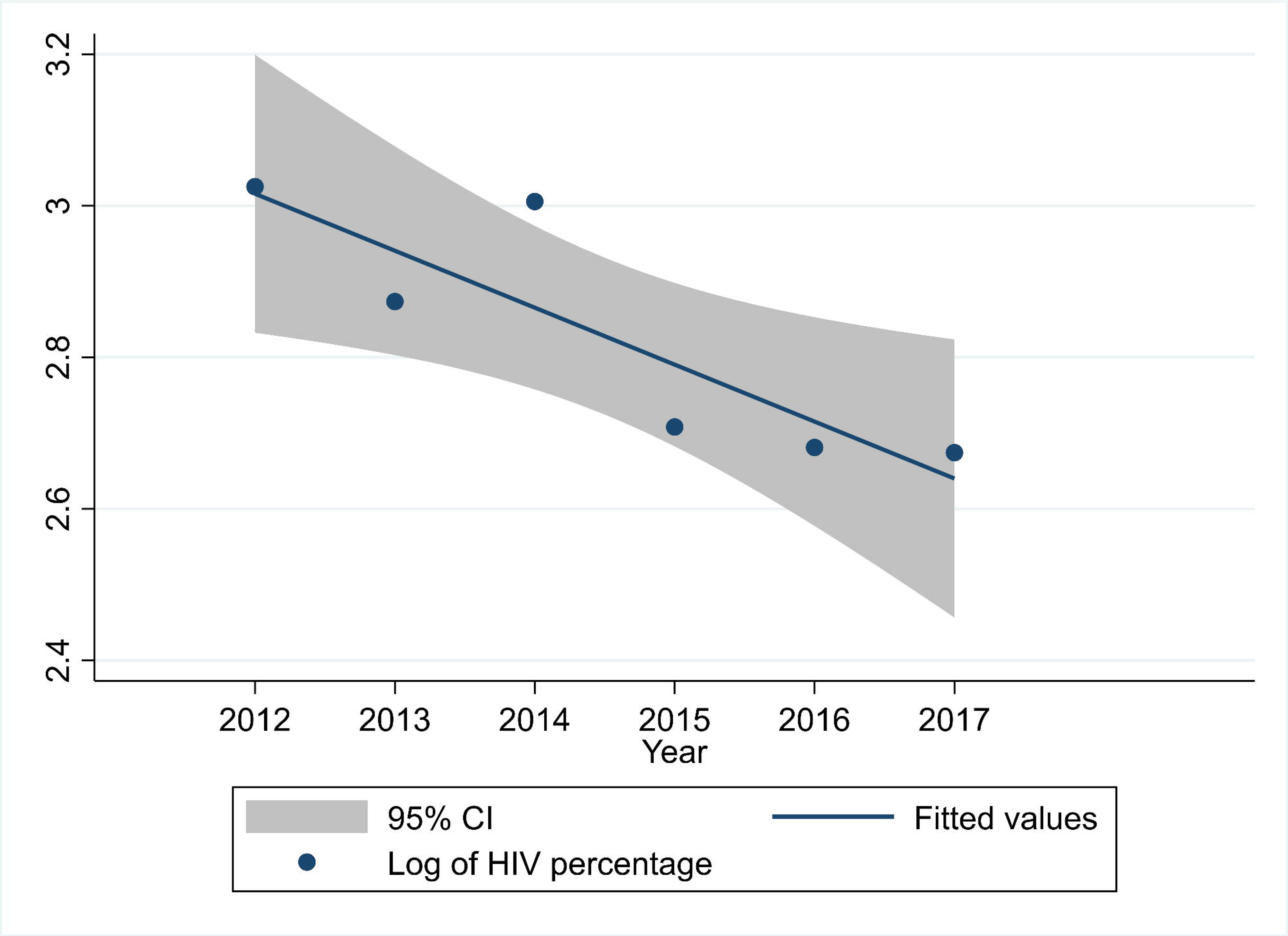
Average annual rate of reduction in the percentage of children with severe acute malnutrition who had HIV in nutritional rehabilitation units between 2012 and 2017. HIV: human immunodeficiency virus

There were no trends in the percentage of males and females from 2011 to 2018 in OTPs (p=0.08) with an AARR of -0.5% (95% CI: -1.0%, 0.1%) (Figure 9, Figure 10A) or NRUs (p=0.3) with an AARR of 0.4% (95% CI: -0.6%, 1.3%) (Figure 9, Figure 10B).

**Figure 9.**
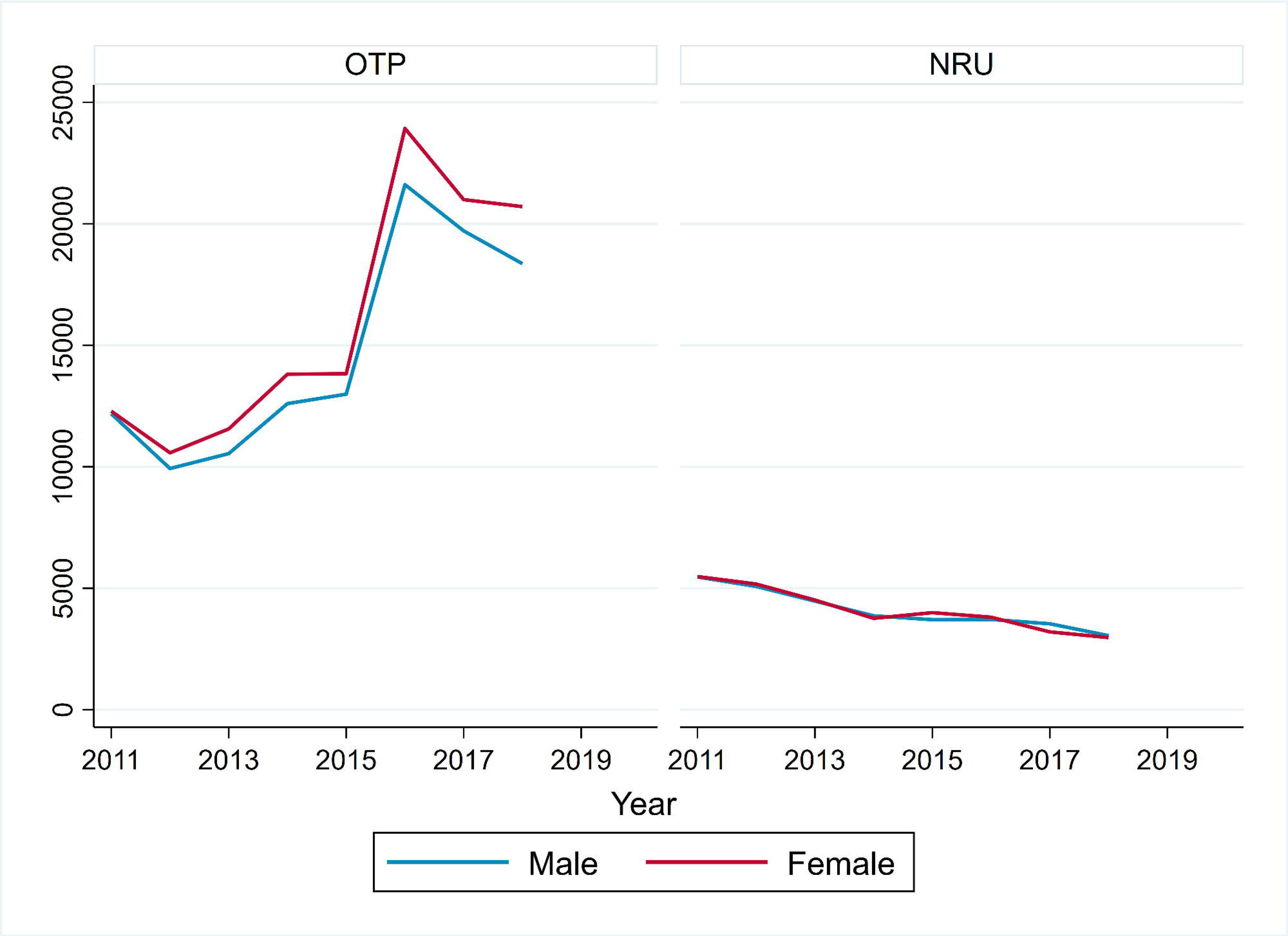
**Trends in the percentage of female children with severe acute malnutrition in outpatient therapeutic feeding programs (A) and nutritional rehabilitation units (B) between 2012 and 2018.**

**Figure 10A.**
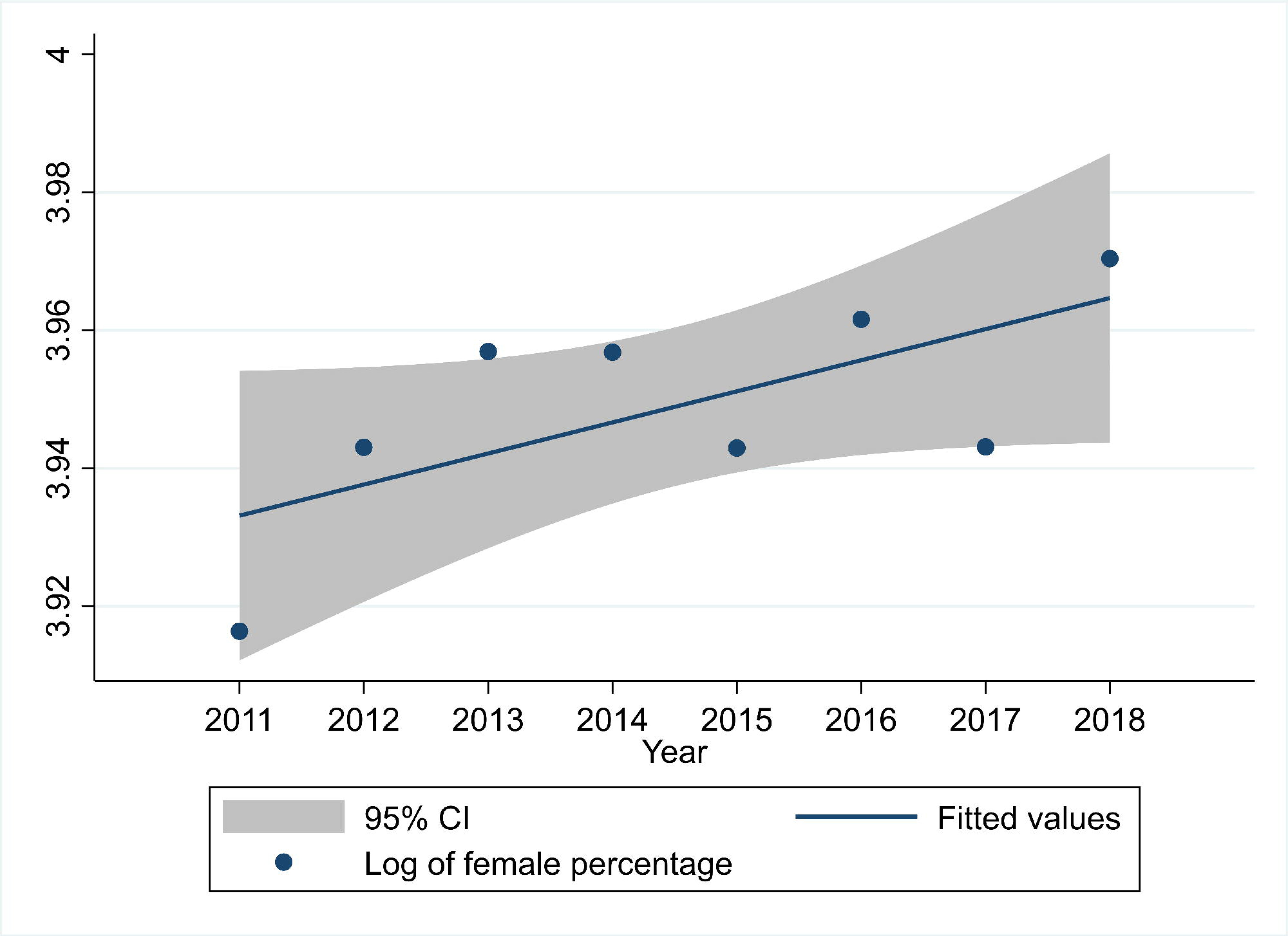
**Average annual rate of reduction in the percentage of female children with severe acute malnutrition in outpatient therapeutic feeding programs between 2012 and 2018.**

**Figure 10B.**
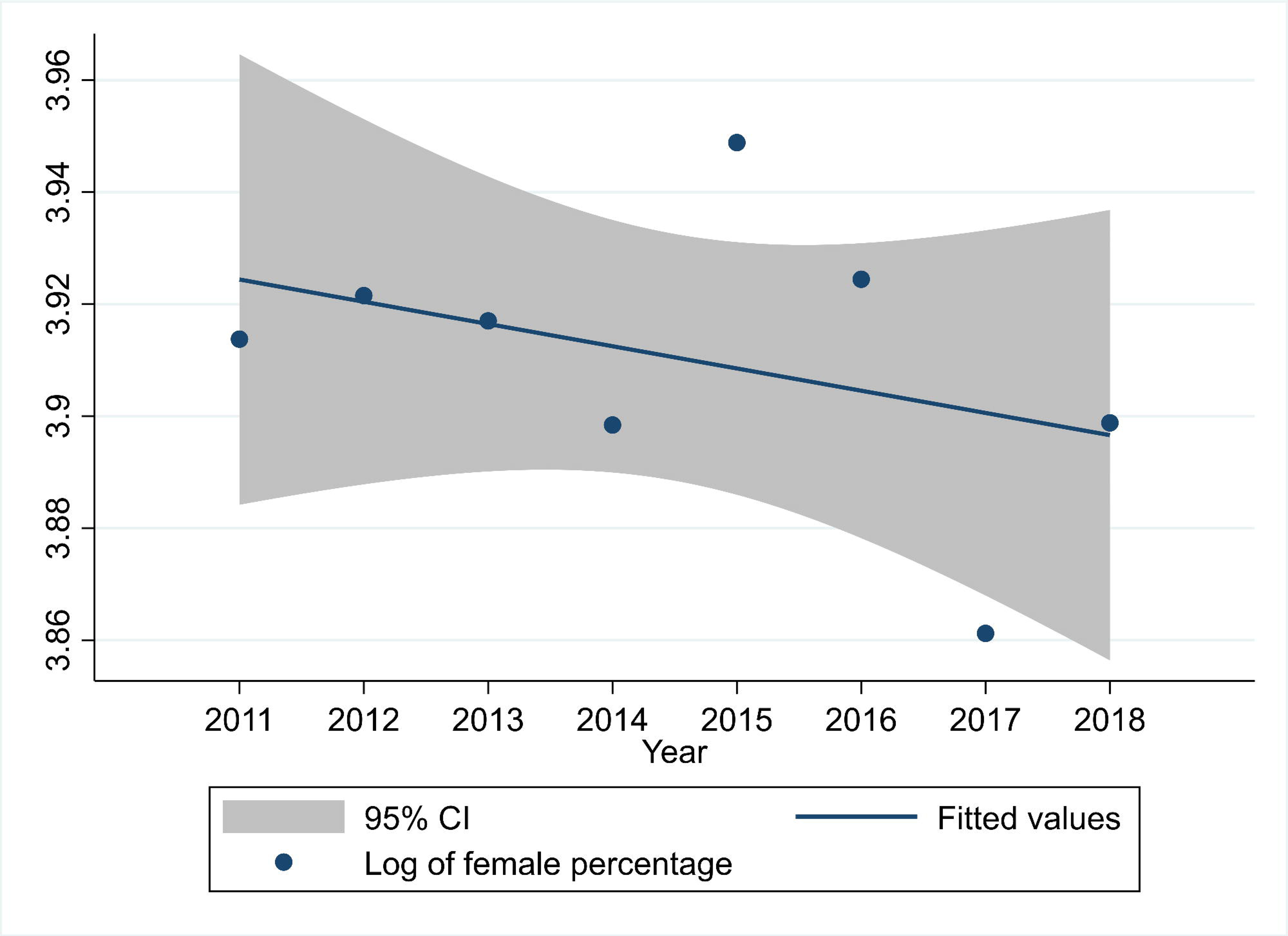
**Average annual rate of reduction in the percentage of female children with severe acute malnutrition in nutritional rehabilitation units between 2012 and 2018.**

### Treatment outcomes in children with severe acute malnutrition

There was no trend in the absolute number of SAM deaths in OTPs (p=0.5) (Figure 11). However, there was a downward trend in death rates in OTP from 1.9% in 2011 to 1.2% in 2019 (p=0.02) with an AARR of 6.0% (95% CI: 1.9%, 10.1%) (Table 3, Figure 12, Figure 13A). Absolute deaths in NRUs decreased from 1167 in 2011 to 594 in 2019 (p=0.01) (Figure 12), yet there was no change in rates of mortality in NRUs (p=0.4) with an AARR of 0.9% (95% CI: - 2.0%, 3.7%) (Table 4, Figure 12, Figure 13B).

**Figure 11.**
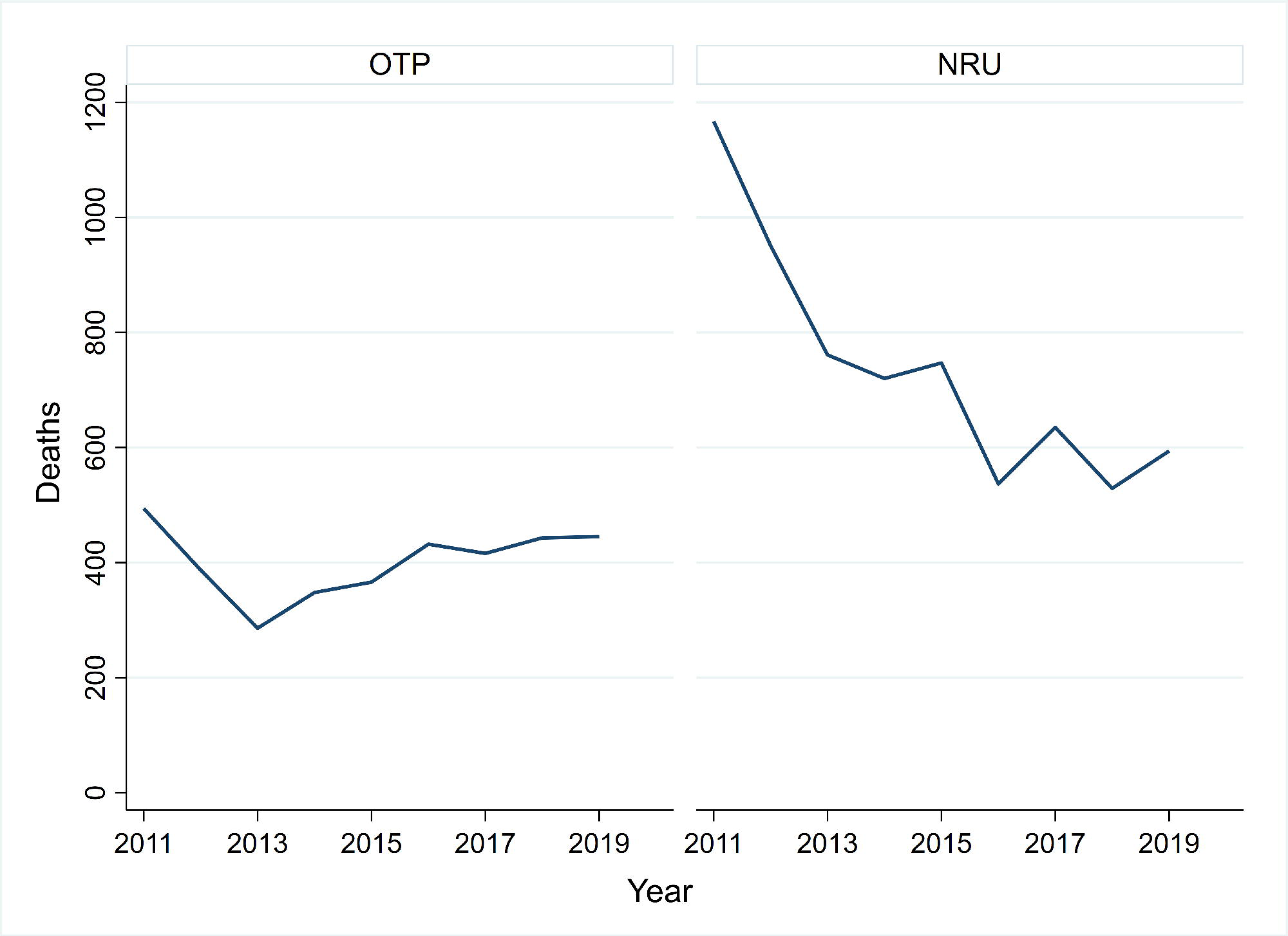
Deaths disaggregated by outpatient therapeutic feeding programs and nutritional rehabilitation units between 2011 and 2019. NRU: nutritional rehabilitation unit; OTP: outpatient therapeutic feeding program.

**Figure 12.**
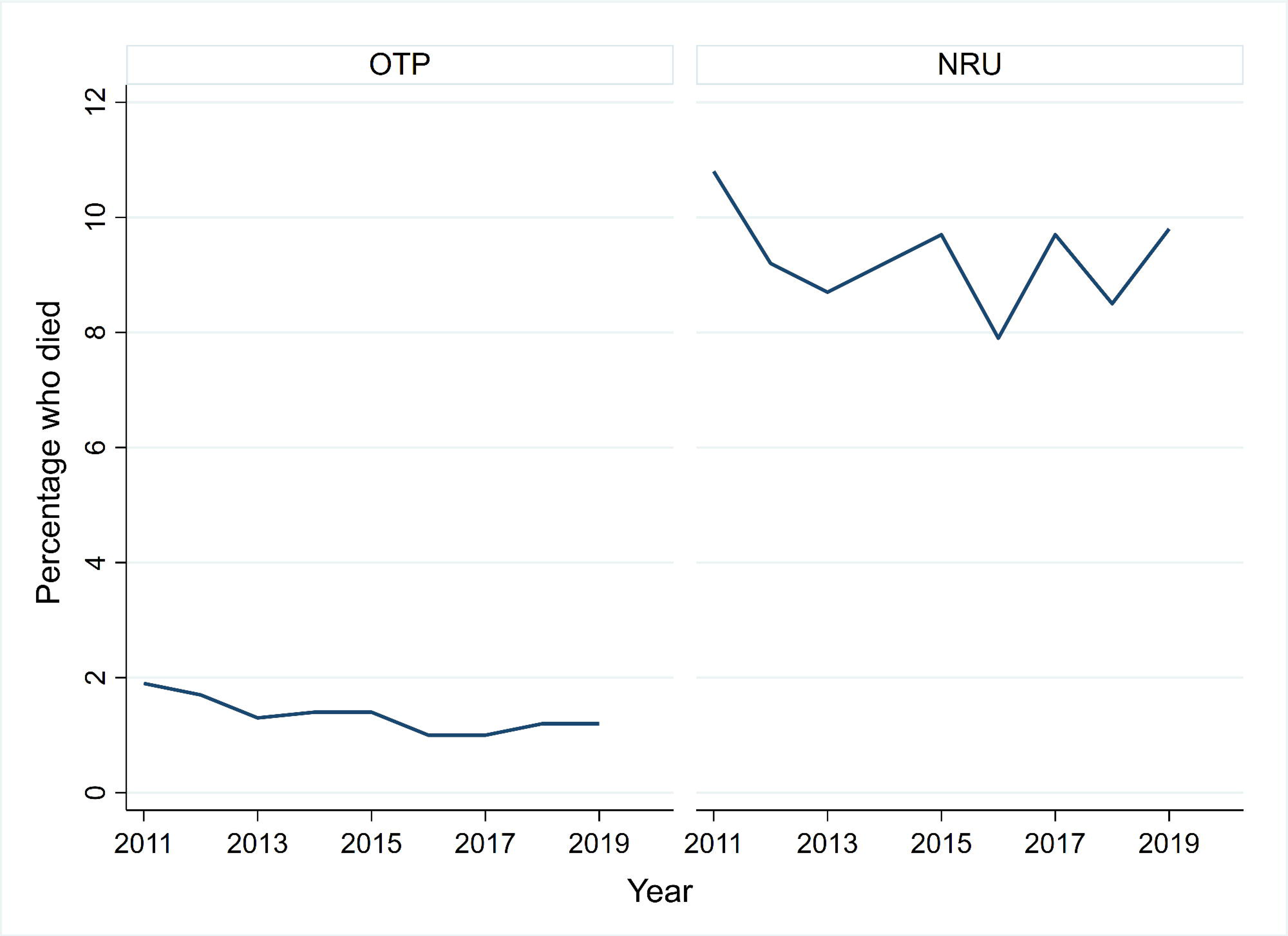
**Trends in the mortality rates in children with severe acute malnutrition in outpatient therapeutic feeding programs (A) and nutritional rehabilitation units (B) between 2011 and 2019.**

**Figure 13A.**
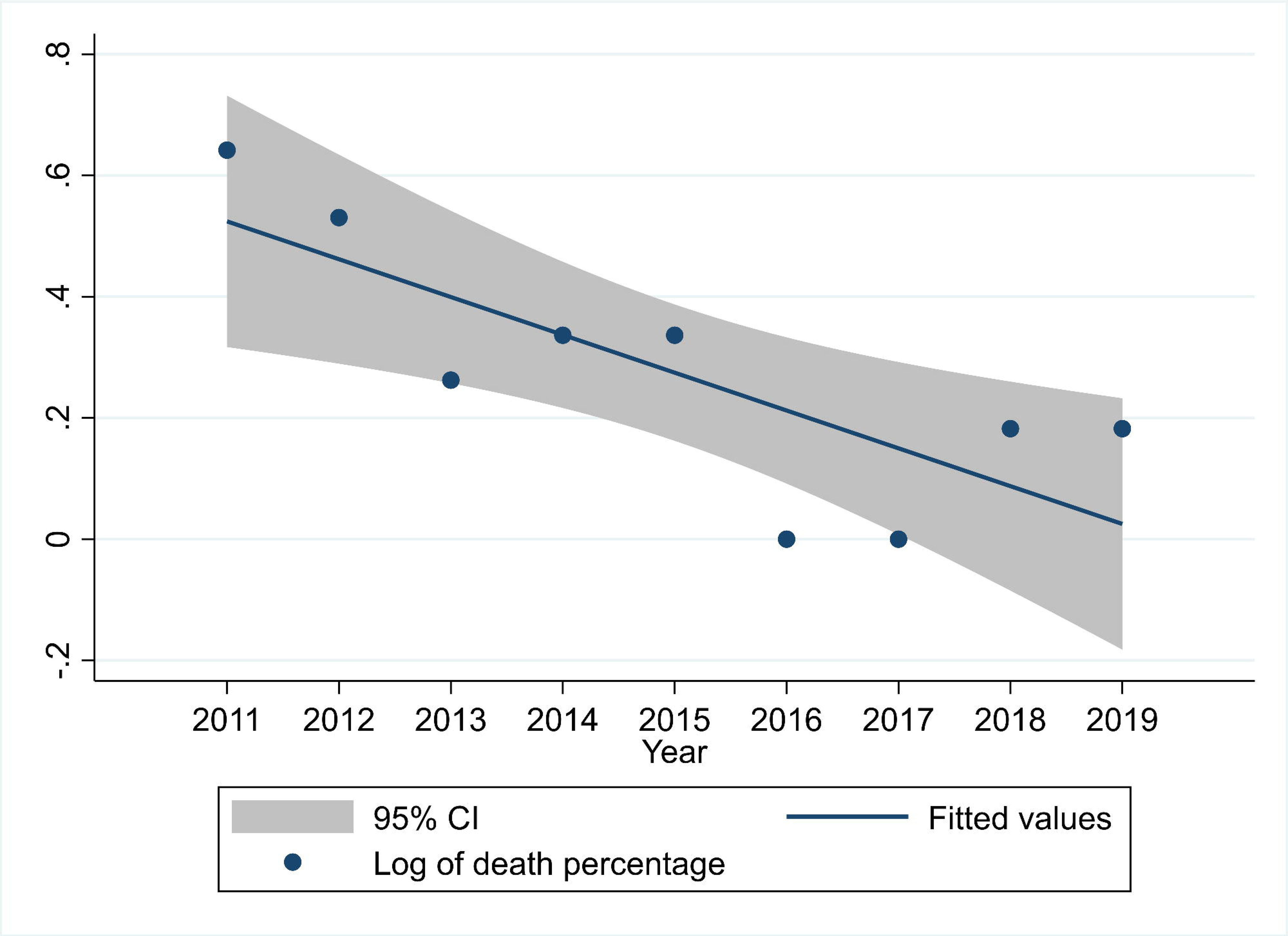
**Average annual rate of reduction in the percentage of children with severe acute malnutrition who died in outpatient therapeutic feeding programs between 2011 and 2019.**

**Figure 13B.**
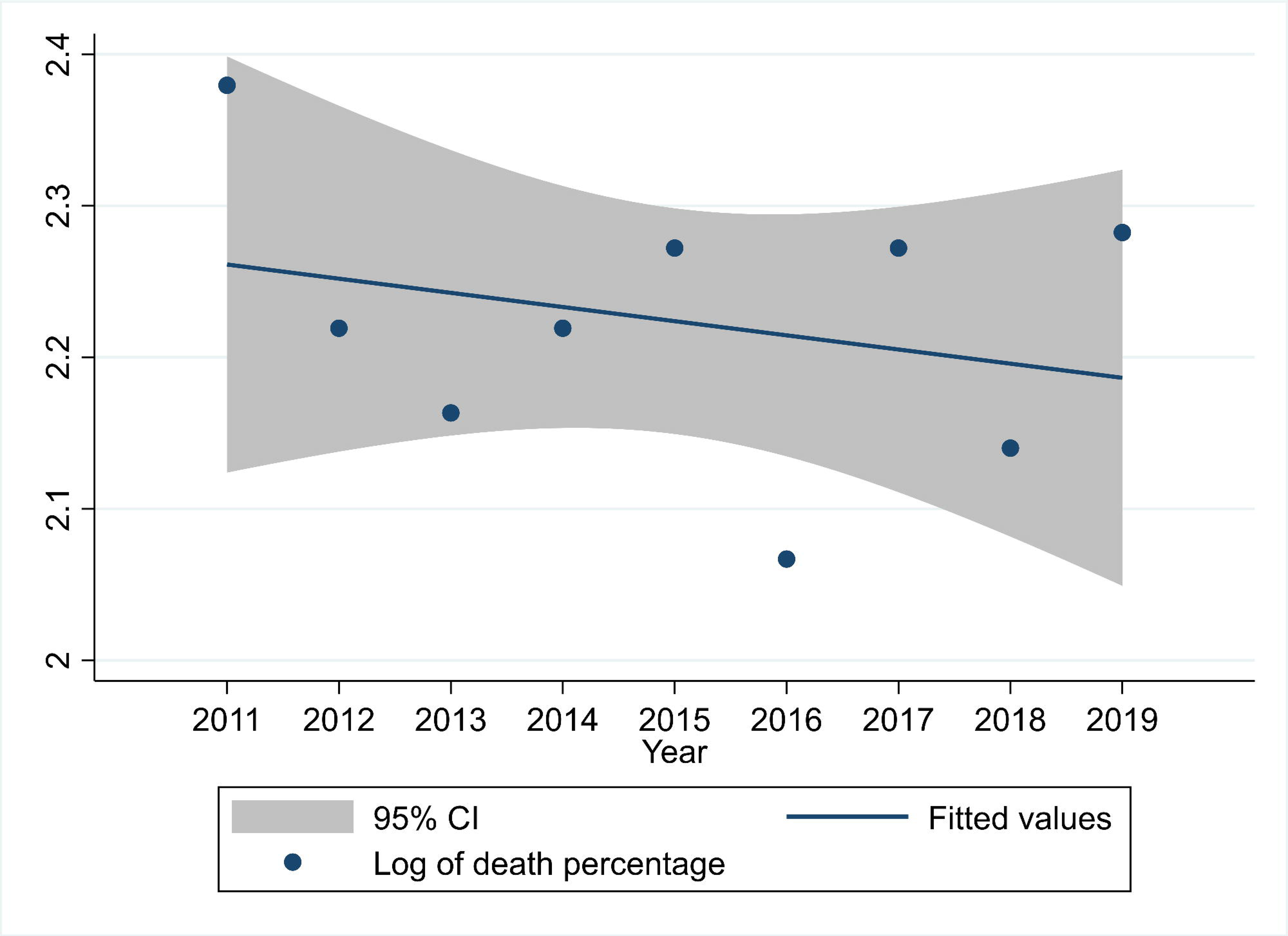
**Average annual rate of reduction in the percentage of children with severe acute malnutrition who died in nutritional rehabilitation units between 2011 and 2019.**

**Table 3.**
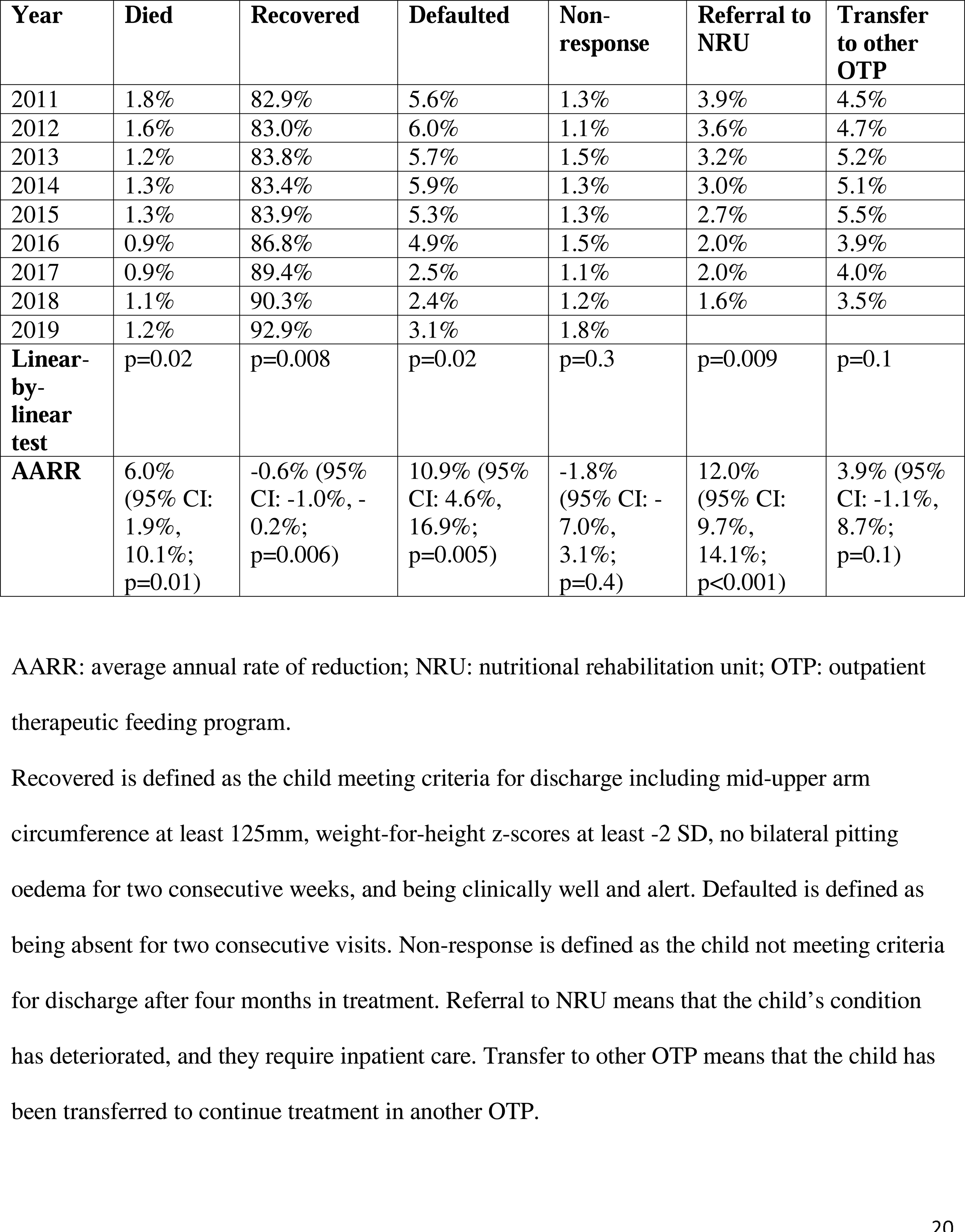
Treatment outcomes at outpatient therapeutic feeding programs between 2011 and 2019.

| Year | Died | Recovered | Defaulted | Non-response | Referral to NRU | Transfer to other OTP |
| --- | --- | --- | --- | --- | --- | --- |
| 2011 | 1.8% | 82.9% | 5.6% | 1.3% | 3.9% | 4.5% |
| 2012 | 1.6% | 83.0% | 6.0% | 1.1% | 3.6% | 4.7% |
| 2013 | 1.2% | 83.8% | 5.7% | 1.5% | 3.2% | 5.2% |
| 2014 | 1.3% | 83.4% | 5.9% | 1.3% | 3.0% | 5.1% |
| 2015 | 1.3% | 83.9% | 5.3% | 1.3% | 2.7% | 5.5% |
| 2016 | 0.9% | 86.8% | 4.9% | 1.5% | 2.0% | 3.9% |
| 2017 | 0.9% | 89.4% | 2.5% | 1.1% | 2.0% | 4.0% |
| 2018 | 1.1% | 90.3% | 2.4% | 1.2% | 1.6% | 3.5% |
| 2019 | 1.2% | 92.9% | 3.1% | 1.8% |  |  |
| **Linear-by-linear test** | p=0.02 | p=0.008 | p=0.02 | p=0.3 | p=0.009 | p=0.1 |
| **AARR** | 6.0% (95% CI: 1.9%, 10.1%; p=0.01) | -0.6% (95% CI: -1.0%, -0.2%; p=0.006) | 10.9% (95% CI: 4.6%, 16.9%; p=0.005) | -1.8% (95% CI: -7.0%, 3.1%; p=0.4) | 12.0% (95% CI: 9.7%, 14.1%; p<0.001) | 3.9% (95% CI: -1.1%, 8.7%; p=0.1) |

AARR: average annual rate of reduction; NRU: nutritional rehabilitation unit; OTP: outpatient therapeutic feeding program.
Recovered is defined as the child meeting criteria for discharge including mid-upper arm circumference at least 125mm, weight-for-height z-scores at least -2 SD, no bilateral pitting oedema for two consecutive weeks, and being clinically well and alert. Defaulted is defined as being absent for two consecutive visits. Non-response is defined as the child not meeting criteria for discharge after four months in treatment. Referral to NRU means that the child's condition has deteriorated, and they require inpatient care. Transfer to other OTP means that the child has been transferred to continue treatment in another OTP.

**Table 4.** Treatment outcomes at nutritional rehabilitation units between 2011 and 2019.

| Year | Died | Stabilized to OTP | Recovered | Defaulted | Medical transfer |
| --- | --- | --- | --- | --- | --- |
| 2011 | 10.8% | 63.1% | 18.7% | 2.7% | 4.7% |
| 2012 | 9.2% | 72.3% | 12.3% | 2.7% | 3.5% |
| 2013 | 8.7% | 74.3% | 10.5% | 2.4% | 4.6% |
| 2014 | 9.2% | 72.3% | 12.2% | 2.7% | 4.4% |
| 2015 | 9.7% | 71.5% | 12.4% | 2.2% | 4.3% |
| 2016 | 7.9% | 74.9% | 11.0% | 2.6% | 3.7% |
| 2017 | 9.7% | 74.9% | 8.8% | 2.4% | 4.1% |
| 2018 | 8.5% | 69.1% | 11.5% | 2.2% | 3.5% |
| 2019 | 11.0% | 80.1% | 13.9% | 2.9% |  |
| **Linear-by-linear test** | p=0.4 | p=0.09 | p=0.2 | p=0.7 | p=0.2 |
| **AARR** | 0.9% (95% CI: -2.0%, 3.7%; p=0.5) | -1.5% (95% CI: -3.2%, 0.3%; p=0.08) | 3.0% (95% CI: -3.1%, 8.8%; p=0.3) | 0.6% (95% CI: -2.6%, 3.7%, p=0.7) | 2.3% (95% CI: -2.0%, 6.4%; p=0.2) |

AARR: average annual rate of reduction; OTP: outpatient therapeutic feeding program.
Stabilized to OTP means that the child's health has stabilized, and they are referred to OTP to
continue treatment. Recovered is defined as recovering in the NRU and the child meeting criteria
for discharge including mid-upper arm circumference at least 125mm, weight-for-height z-scores
at least -2 SD, no bilateral pitting oedema for two consecutive weeks, and being clinically well
and alert. Defaulted is defined as being absent for two consecutive days. Non-response is defined
as not meeting criteria for discharge after four months in treatment. Medical transfer means the
child is transferred to another inpatient facility for further medical investigation and treatment.

There was an increase in recovery percentage from 82.9% in 2011 to 92.9% in 2019 in OTPs (p=0.008) with an AARR of -0.6% (95% CI: -1.0%, -0.2%). There was a decrease in percentage of children who defaulted from OTPs from 5.6% in 2011 to 3.1% in 2019 (p=0.02) AARR of 10.9% (95% CI: 4.6%, 16.9%). There was no change in non-response (p=0.3) in children with SAM at OTPs with an AARR of -1.8% (95% CI: -7.0%, 3.1%). There was a decline in the percentage of children who were referred from OTP to NRUs from 3.9% in 2011 to 1.6% in 2018 (p=0.009) with an AARR of 12.0% (95% CI: 9.7%, 14.1%) but no difference in transfer to OTP (p=0.1) with an AARR of 3.9% (95% CI: -1.1%, 8.7%) (Table 4).

There were no differences in other treatment outcomes in children with SAM who were admitted to NRUs between 2011 and 2019, including stabilized to OTP (p=0.09) with an AARR of -1.5% (95% CI: -3.2%, 0.3%), recovered (p=0.2) with an AARR of 3.0% (95% CI: -3.1%, 8.8%), defaulted (p=0.7) with an AARR of 0.6% (95% CI: -2.6%, 3.7%), and medical transfer in NRUs (p=0.2) with an AARR of 2.3% (95% CI: -2.0%, 6.4%) (Table 4).

## Discussion

This is the first analysis of national trends in SAM admissions, characteristics, and treatment outcomes over the course of a decade in any country that has implemented CMAM. These data from Malawi show that there was an increase in SFP and OTP admissions and a major reduction in NRU admissions between 2011 and 2019, with mortality in NRUs being high at between 7.9% and 11.0%. Admissions for SAM in both OTPs and NRUs were generally higher in the first few months of each year.

The percentage of children with SAM who had oedema compared to severe wasting declined in OTPs and NRUs across Malawi within the time examined. The primary reason for this is likely to be that Malawi introduced highly effective active case finding in the community, which could identify more children with wasting who may have previously been missed. On the other hand, the drop in the percentage of children with oedema compared to severe wasting does not necessarily represent a change in the phenotype.

HIV rates in children with SAM were seen to be lower in both CMAM settings over time, albeit non-significant in OTPs. This may represent a shift in the characteristics of children with SAM in Malawi particularly in NRUs. HIV is known to been associated with elevated risk of mortality in children admitted for inpatient treatment of SAM (Bandsma et al., 2019; Heikens et al., 2008; Rytter et al., 2017; Trehan et al., 2012), but the data indicated that SAM mortality in NRUs has not declined. However, the CMAM data available did not link HIV and mortality, which makes it impossible to determine whether this is the case from this analysis.

Though a fraction of total admissions for SAM is to NRUs, the absolute number of deaths remains higher in NRUs than in OTPs, with 445 deaths recorded in OTPs and 594 in NRUs in 2019. The Sphere Handbook, which Malawi’s CMAM reporting system follows, states that mortality rates should be below 10% for combined OTP and NRU outcomes (Sphere Association, 2018). Malawi has been near this threshold in NRUs, with mortality rates of 9.8% in 2019 with four months of that year in which NRUs exceeded mortality rates of 10%. Evidently, children admitted for inpatient treatment of SAM remain exceptionally vulnerable even though there are fewer children being treated in NRUs.

Results from this analysis of CMAM trends in Malawi point towards key actions to be taken. The first set of actions is to further support and strengthen active case finding and early identification of wasting, augmenting treatment in community settings where most children with wasting can be managed, and improving the referral system from OTP to NRUs for children with clinical complications. Additionally, the quality of care of children admitted to NRUs should be enhanced considering the stagnant mortality rates. A recent implementation evaluation showed that a 17-month quality improvement initiative beginning in April 2016 at seven hospitals in Malawi with SAM mortality rates above 10% improved the assessment of clinical complications and nutritional status, prevention and treatment of dehydration, and immediate cautious feeding of children with SAM (Kauchali et al., 2022). However, the mortality rates remained over 10%, with death audits showing that deaths were often attributed to delayed presentation, clinical complications, and inability to access antibiotics. The authors suggested post-training support for healthcare workers at NRUs, integration with emergency care in alignment with Emergency Triage Assessment and Treatment (ETAT), and pre-service training of healthcare workers which started in Malawi in 2016 (Kauchali et al., 2022). With fewer children admitted to NRUs meaning that many are not at capacity (Daniel et al., 2019; Kouam, 2016) – and many sitting empty – there is strong potential for this to be done without greatly increasing resource requirements.

Importantly, there are several key limitations of this evaluation. One is that patterns observed across NRUs and OTP may not be representative of all districts in Malawi. Findings were not examined by district due to unavailability of these disaggregated data. Furthermore, annual data are presented but there is potential for issues with regards to data quality and reporting that may impact the findings. There are also no data on treatment coverage which makes it challenging to draw conclusions on admissions in particular. Future programmatic data should include details on coverage and be presented by district and specific NRU and OTP sites if possible. Additional limitations are that the data do not allow for exploration of direct relationships between admissions, characteristics, and treatment outcomes in children with wasting. The data are also not disaggregated by age, including infants under six months compared to infants and children over six months. In summary, there are many ways to improve the usability of programmatic data moving forward, which should be coupled with improving quality and completeness of these data to get a deeper understanding of these trends over time.

## Conclusions

There has been an increase in OTP admissions of children with SAM since the implementation of CMAM and a corresponding significant decline in NRU admissions in Malawi. These trends in NRU admissions as well as decreasing percentage of children with SAM who have oedema demonstrate the positive impact of active case finding, particularly to identify wasting. However, the mortality rate for children admitted to NRUs has not changed. These findings signal the importance of reinforcing case finding to identify wasting, effective treatment of children who have wasting in community settings, timely referral to NRUs for children with SAM and clinical complications, and improving quality of care at NRUs for those at highest risk.

## Data Availability

All data are available upon reasonable request to the authors.

## Acknowledgements

We would like to sincerely thank the Malawi Ministry of Health for sharing these data to complete this analysis, including staff members involved in collecting these data at sites across Malawi over the years.

## Author contributions

AID, SK, CM, WV, RB, and IP contributed to the conception and design of this analysis. AID performed the statistical analysis and drafted the manuscript. All authors contributed to the interpretation of the data. All authors were involved in critical review of the manuscript and approved the final manuscript.

